# Leveraging long-term smartwatch data to inform Parkinson’s disease progression, subtypes, and risk

**DOI:** 10.1101/2023.09.13.23295404

**Authors:** Ann-Kathrin Schalkamp, Kathryn J Peall, Neil A Harrison, Valentina Escott-Price, Cynthia Sandor

## Abstract

Use of digital sensors to passively collect long-term, longitudinal data offers a step change in our ability to monitor Parkinson’s disease (PD). However, to date the evaluation of long-term digital sensor data has been neglected in favour of evaluating short-term data collected in controlled settings. To address this, we combined longitudinal clinical and biological assessment data from the Parkinson’s Progression Marker Initiative (PPMI) cohort with long-term (mean: 485 days) at-home digital monitoring data collected with the Verily Study Watch. We then derived digital timeseries components leveraging the long-term monitoring of the PPMI. We found three key findings: Firstly, that these digital timeseries components correlated with the rate of progression of motor (r = 0.23, p-value = 8.5×10^-3^, r = 0.26, p-value = 2.2×10^-3^) and autonomic symptoms (r = −0.23, p-value = 8.2×10^-3^), impairments in daily living (r = 0.26, p-value = 2.5×10^-3^), increase in medication requirements and complications (r = −0.25, p-value = 4.2×10^-3^), and rate of increase in cerebrospinal fluid (CSF) tau (ptau: r = 0.28, p-value = 2.6×10^-3^; ttau: r = 0.34, p-value = 1.2×10^-4^). Second, we derived digitally informed subtypes of PD and found higher similarity with CSF (0.35) and DaTscan (0.35) subtypes than has been found for previously published subtypes (CSF: 0.31±0.01, DaTscan: 0.31±0.02). Finally, we showed that long-term digital monitoring can inform PD risk and sensitively detect individuals with probable prodromal PD. Our findings highlight the wealth of application areas for digital sensors in PD research.

## Introduction

Traditionally, Parkinson’s disease (PD) monitoring has relied on periodic clinical assessments to track disease progression and inform changes in clinical management. Passive patient monitoring approaches via digital health technologies (DHTs) that can continuously track physical activity, sleep, and vital signs could provide convenient, cost-effective, and more comprehensive information of an individual’s health status.

Despite one of the main advantages of digital sensors being their ability to passively collect data over long periods in real-world settings, most research to date has focussed on short periods of data collected in controlled environments. For example, long-term data collected in free-living conditions has the potential to complement or even better describe the heterogeneous disease progression observed in PD, which is currently evaluated using the Unified Parkinson’s Disease Rating Scale (UPDRS) recorded at each clinic consultation. Digital sensors can provide a more objective measure of sleeping behaviour, physical activity, and vital signs, that is potentially more closely related to disease pathophysiology. Thus, another area where passively collected digital data could play a role is in identifying PD subtypes aiding the stratification of PD. This is an active area of research with a common consensus yet to be achieved. The final area where digital data could be useful is in providing prognostic information and risk stratification benefitted by the ability of long-term digital device data to capture subtle motor symptoms during early disease stages.

Here, we used multi-modal data including smartwatch data, biological measures (namely from Dopamine Transporter Scan (DaTscan) and Cerebrospinal Fluid (CSF)), medication use, and standardised questionnaires and tests detailing motor, cognitive, autonomic, psychiatric, and daily living impairment from the Parkinson’s disease Progression Marker Initiative (PPMI) (Marek et al., 2018) to investigate how long-term collected smart watch data may contribute to prognosis and stratification. More specifically, we extracted informative features from the digital timeseries data, using these to 1) determine their relationship to clinical and biological progression; 2) extract digital subtypes, comparing these to known clinically and biologically informed subtypes; 3) derive a digital risk score for PD and relate it to a known prodromal risk score.

## Results

### Digital markers differ between PD cases and controls

At the time of data retrieval (November 2022), the PPMI dataset provided a mean of 485 days of at home monitoring for 14 features describing physical activity (step count, walking minutes), sleep (total time, REM time, NREM time, deep NREM time, light NREM time, wake after sleep onset (WASO), awakenings, sleep efficiency), and vital signs (pulse rate, mean root mean squared successive differences (RMSSD) (heart beat), median RMSSD, RMSSD variance) in 1-hour intervals (Supplemental Table 1) for 343 subjects derived from the multi-sensor Verily Study Watch. The cohort included individuals diagnosed with PD (n = 149), individuals identified based on specific genetic and/or prodromal markers, the prodromal group (n = 158), and unaffected controls (n = 35). Seven individuals originally assigned to the prodromal group received a diagnosis of PD after recruitment. Six of those received the diagnosis before smartwatch data collection, for one the date of phenoconversion was unknown. These were removed from the prodromal group and analysed separately.

Overall, individuals diagnosed with PD showed significant impairments for six of 14 digital markers compared to the unaffected controls and the prodromal group (Supplemental Figure 1, Supplemental Table 2). To assess which markers were generally indicative of PD, we computed the mean over the whole observation time. This showed that all physical activity markers were reduced in PD compared to controls: step count (p-value = 5.08×10^-10^), walking minutes (p-value = 1.13×10^-7^). Four of the eight sleep markers were also significantly lower in individuals diagnosed with PD compared to controls; sleep length (p-value = 1.46×10^-3^), sleep efficiency (p-value = 9.6×10^-9^), REM (p-value = 2.34×10^-9^), and deep NREM sleep length (p-value = 1.87×10^-6^). None of the four digital vital signs were significantly different between PD cases and healthy controls. The differences in physical activity and sleep markers were also observed between the PD and the prodromal group. More specifically, the prodromal and control groups were comparable across all measures except for step count, which was significantly lower in the prodromal group (p-value = 1.12×10^-2^). Further, compared to the PD cases, the prodromal group had reduced pulse rate (p-value = 2.82×10^-5^), fewer awakenings (p-value = 2.61×10^-3^) and WASO (p-value = 1.21×10^-3^), and more NREM sleep time (p-value = 1.89×10^-3^). These differences were not present when comparing healthy controls to the PD cases. The prodromal group was heterogeneous with some individuals being carriers of known genetic mutations (*GBA, LRRK2, SNCA, Parkin, Pink1*) and others displaying prodromal symptoms (hyposmia, RBD). We therefore split the prodromal group into subgroups based on the genetic risk and prodromal markers and found that the prodromal *GBA* variant group (N = 90) showed significant differences to the PD cases in pulse rate (p-value = 2.12×10^-4^) and WASO (p-value = 9.53×10^-4^) (Supplemental Figure 2, Supplemental Table 3). These were not found for any other prodromal group nor the healthy controls when compared to the PD cases.

### Clinical and biomarker indices of progression are reflected in the digital timeseries data

We used tsfresh for an automatic extraction of 783 features for each subject for each of the digital timeseries over the observation period (2018-2022), leading to a total of 10962 features per subject (Figure 1). For details see the Methods section, in brief, we used Principal Component Analysis (PCA) to compress all extracted features, then investigated the first ten principal components, from here on referred to as the digital timeseries components. The clinical and biological data (2010-2021) was used to derive estimates for 1) the severity at diagnosis and 2) the rate of progression using linear mixed effects models. Composite scores for specific domains, namely motor, cognitive, psychiatric, autonomic, daily living, medication, and DaTscan striatal binding ratio (SBR) and DaTscan asymmetry were obtained via PCA (see Methods for details). We further evaluated CSF GFAP, NFL, ttau, ptau, Amyloidbeta, and alpha-synuclein.

**Figure 1:**
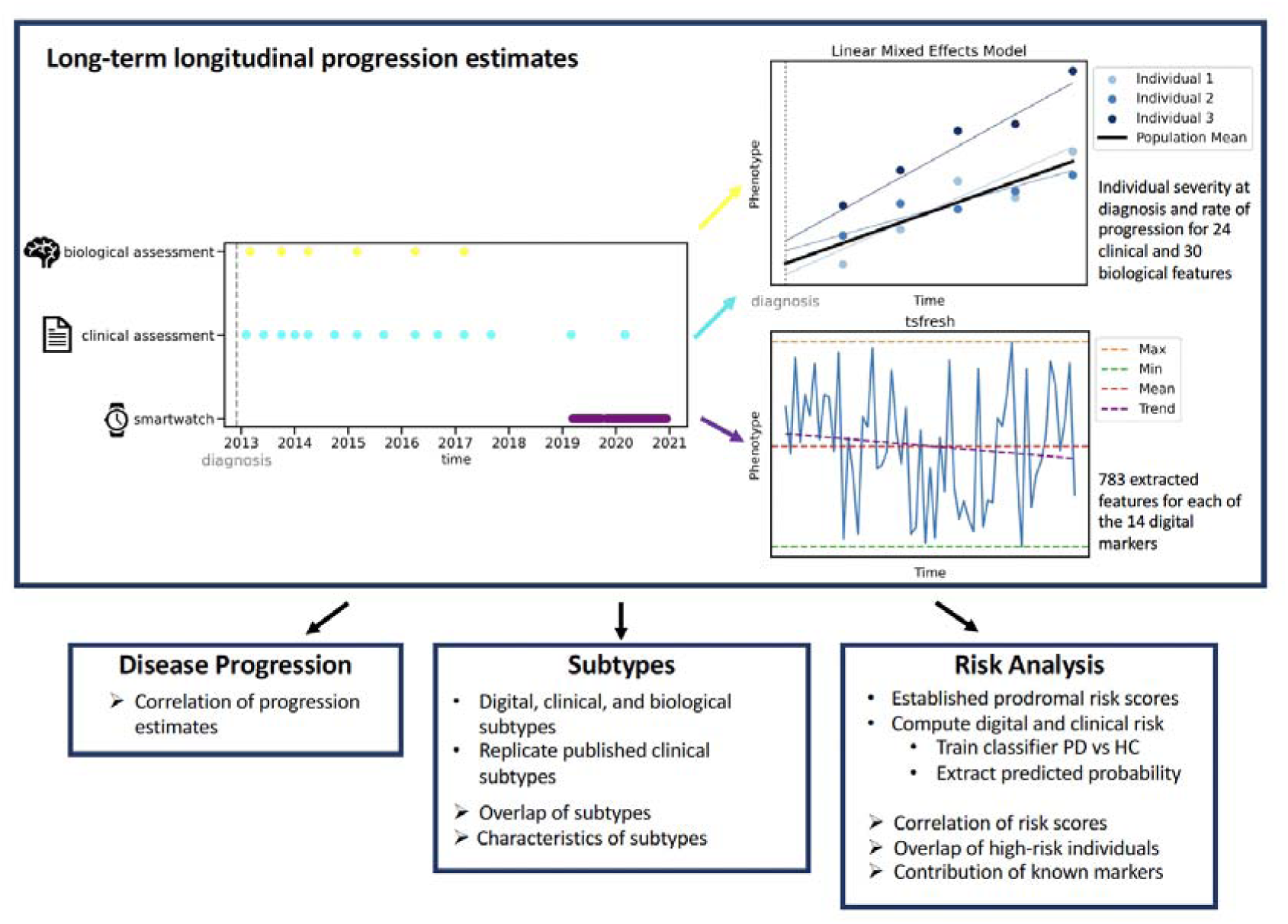
Overview of methods. The longitudinally collected data was processed to obtain progression estimates for clinical and biological data via linear mixed models and for smartwatch data via tsfresh. The extracted estimates were leveraged to inform disease progression, subtypes, and risk analyses.

None of the digital timeseries components showed a significant correlation with the severity of any of the clinical signs and biological markers at the time of diagnosis, potentially due to the >10-year time gap between events (Supplemental Table 4). However, the digital timeseries components did significantly correlate with *rate of progression* (i.e. change) of several clinical and biological markers: motor signs (digital timeseries component 5: r = 0.23, p-value = 8.5×10^-3^, and digital timeseries component 7: r = 0.26, p-value = 2.2×10^-3^), autonomic functioning (digital timeseries component 9: r = −0.23, p-value = 8.2×10^-3^), independence in daily living (digital timeseries component 4: r = 0.26, p-value = 2.5×10^-3^), medication (LEDD, UPDRS III ON – UPDRS III OFF, UPDRS IV) (digital timeseries component 0: r = −0.25, p-value = 4.2×10^-3^), and the biological markers ptau (digital timeseries component 6: r = 0.28, p-value = 2.6×10^-3^) and ttau (digital timeseries component 6: r = 0.34, p-value = 1.2×10^-4^) (Figure 2, Supplemental Table 4).

**Figure 2:**
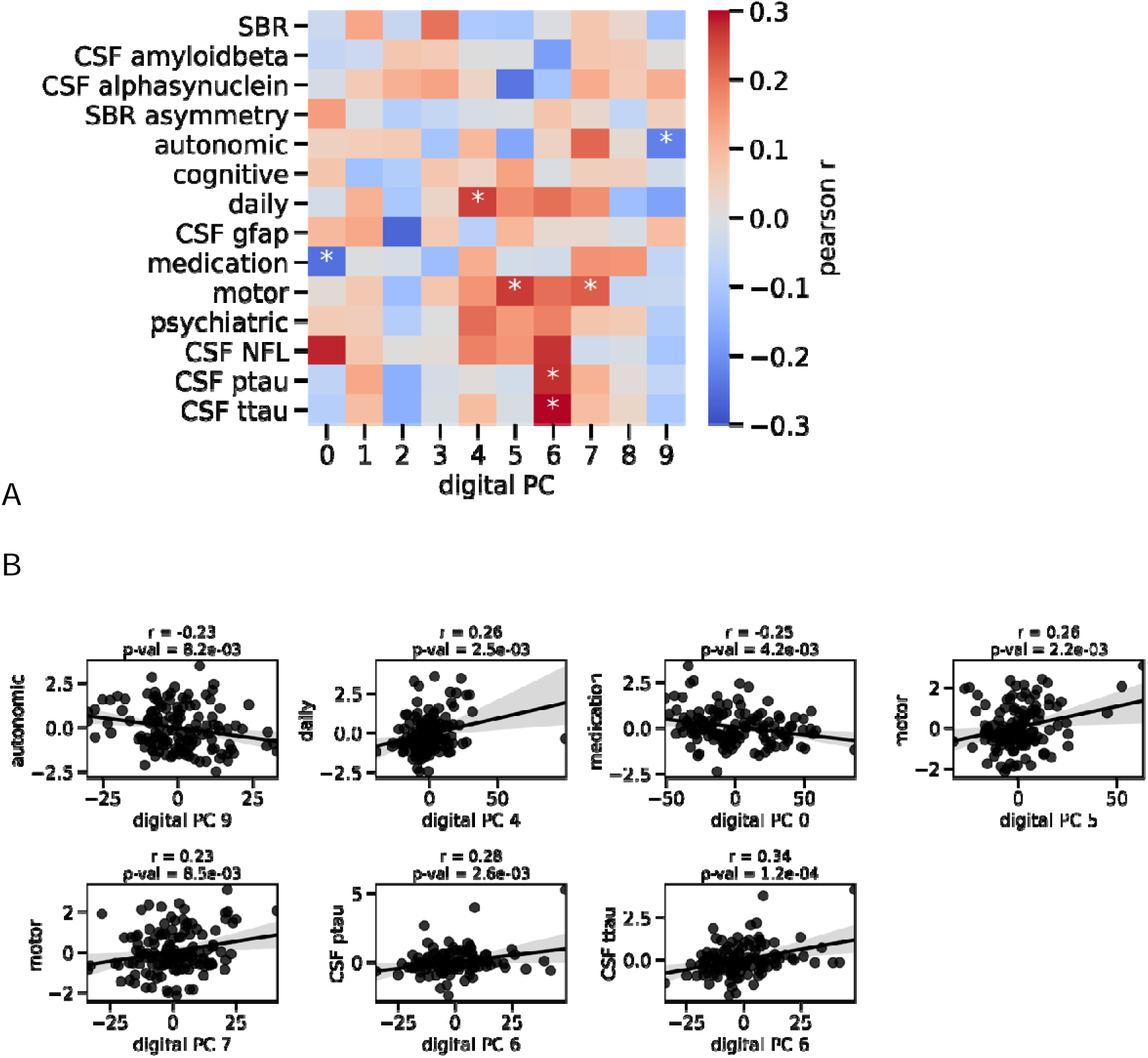
Correlation of digital progression and clinical progression. Associations between the clinical progression and digital timeseries features are shown. (A) The heatmap displays the Pearson’s r coefficient for the first 10 digital progression principal components and the clinical principal components for the progression (slope). Asterisk indicates significant correlation after 0.05 Bonferroni-correction. (B) The scatterplots display the relation between the identified significant pairs. The line indicates the linear fit and the title displays the Pearson’s coefficient and the associated uncorrected p-value.

To investigate this further, we next identified the contributing timeseries features for each of the associated digital timeseries components (Supplemental Figure 3). The digital timeseries features contributing to digital timeseries components five and seven related to motor progression were RMSSD features, the independence in daily living was represented by component four made up of features of mean pulse rate, the component nine correlating with autonomic functioning was made up of REM sleep and physical activity markers, and medication was represented by component 0 including WASO features. The rate of progression of CSF ptau and ttau related to digital timeseries component six representing WASO and NREM sleep time.

Psychiatric and cognitive signs and symptoms, CSF Amyloidbeta, GFAP, NFL, and total alpha-synuclein, and DaTscan progression could not be explained through digital data. Overall, half of the progression aspects of PD could be captured with digital data while the severity at diagnosis could not be captured with the digital data, likely due to the long timespan between diagnosis and digital data collection.

### Digital subtypes are most similar to biologically derived clusters

Several studies have investigated potential subtypes (Dadu et al., 2022; Fereshtehnejad et al., 2017; Lawton et al., 2018; Zhang et al., 2019) of PD based on clinical or biological data, but no consensus has been reached to date. We compared our digital clustering to those previously published subtypes and our own biological and clinical clusters. Compared to the published subtypes, which were either built using (i) a single clinical assessment (Fereshtehnejad et al., 2017), (ii) developed from clinical data captured longitudinally across multiple visits (Dadu et al., 2022), or (iii) used combined clinical and biological timeseries data (Zhang et al., 2019), our digital sub-typing used long-term digital data collected continuously. We further developed our own clinical and biological sub-typings based on the extracted progression estimates (see above). Similarity of clusters were quantified using a composite score of three different metrics of cluster similarity (see Methods) ranging from 0 to 1.

Overall, the clusters in our analysis showed a mean overall similarity of 0.31±0.06 (Supplemental Table 5). The two published clinical sub-typings using longitudinal data (Zhang2019 and Dadu2022) had the highest similarity (overall score = 0.48). Our digital clusters demonstrated a comparable similarity with all previously published subtypes (0.39±0.01, ranging from 0.38 to 0.41) as the published sub-typings had amongst themselves (0.39±0.06, ranging from 0.35 to 0.48). However, the digital clusters had higher a similarity to subtypes derived from progression estimates for CSF (0.35) and DaTscan data (0.35) than all other sub-typings (CSF: 0.31±0.01, ranging from 0.3 to 0.31, DaTscan: 0.31±0.02, ranging from 0.29 to 0.34). This was also true for the clustering by Zhang et al. (2019) despite this using information on CSF and DaTscan in the formation of the clusters. Although the overall consistency between sub-typings remains rather low (<0.48), the results presented here highlight a potentially important link between subtypes from digitally measured phenotypes and subtypes from CSF and DaTscan data.

Each subtype showed primarily significant differences for the domain they were trained on (Figure 3, Supplemental Table 6). Additionally, our own clinical clustering (lmmclinical) demonstrated significant differences in the digital timeseries component 9 representing REM sleep features (FDR corrected p-value = 2.93×10^-2^). The published sub-typings showed significant differences in terms of clinical severity and progression estimates. For Fereshtehnejad et al. (2017) the cluster 0 (mild motor predominant) and 2 (diffuse malignant) displayed significant differences across all clinical progression estimates, with exception of cognition. Although Fereshtehnejad et al. (2017) did not consider longitudinal data in their clustering and we applied their method to the last clinic visit data available, the rate of progression estimates differed significantly between their clusters, as also highlighted in their own manuscript. For the clustering by Dadu et al. (2022), which was trained on longitudinal clinical data, the clusters differed in autonomic, impairment in daily living, and psychiatric progression rates and the clusters showed differences for all severity at diagnosis estimates, except cognition. The longitudinal clustering based on clinical, CSF, and DaTscan data by Zhang et al. (2019) only displayed significant differences between cluster 0 and 2 for autonomic (FDR corrected p-value = 3.34×10^-2^) severity at diagnosis. Of note, no reporting was possible for our digital and lmmclinical cluster 2 as these were only made up of one subject after removal of subjects with incomplete data.

**Figure 3:**
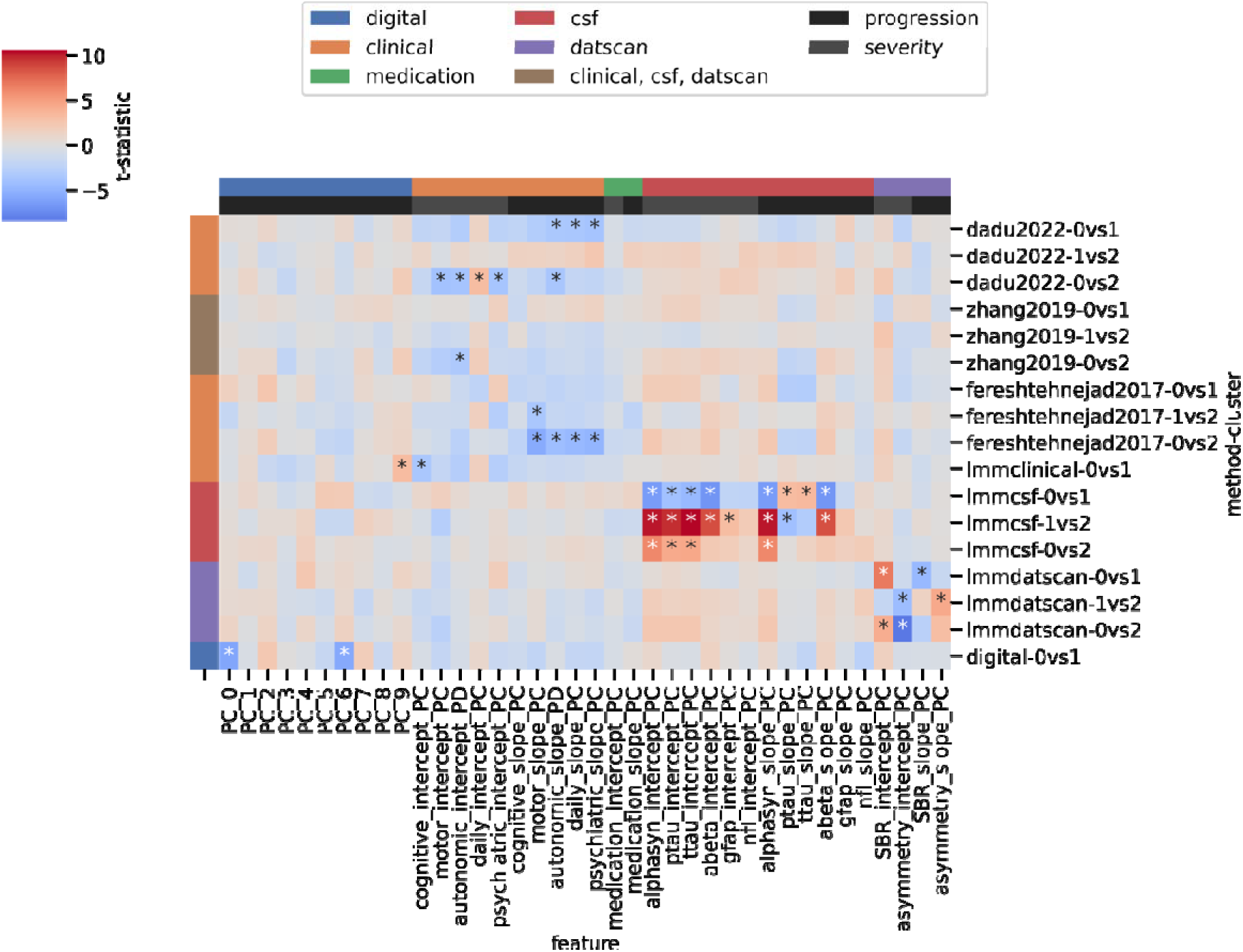
Digital, clinical, and biological severity and rate of progression differences between subtypes. The heatmap displays the t-statistic on a divergent colour gradient ranging from blue (negative correlation) to red (positive correlation) for each severity and progression marker (x-axis) between the subtypes (y-axis). The asterisk signifies 0.05 FDR corrected significance by two-sided T-tests. For each clustering method the differences between each cluster pair are shown for the digital, clinical, medication, CSF, and DaTscan severity and progression estimates. The colours on the left indicate from which data modality the subtypes were derived. The colours on the top identify which modality the examined features belong to.

### Digital risk scores identify individuals with probable prodromal PD as high-risk

We derived risk scores for PD for the digital timeseries data and the last available clinical data respectively by training classifiers to identify PD from healthy controls. Those models were then used to obtain risk scores for the prodromal group. We compared our risk scores to a published risk score based on risk factors and prodromal symptoms (Heinzel et al., 2019).

The predicted risk scores from the digital data were significantly correlated with the prodromal risk scores extracted with the model by Heinzel et al. (2019) (Figure 4, p-value = 1.7×10^-4^) and the clinically derived scores were also associated with the established model (p-value = 5.6×10^-6^). The model by Heinzel et al. (2019) used binary identifiers of which some, for example for subthreshold parkinsonism/UPDRS III > 6, were built using the clinical data (Supplemental Table 7), which were included in the clinical risk model. The digital risk was not significantly correlated with the clinical risk (p-value = 2×10^-1^), possibly indicating a distinct selection of high-risk individuals between the two approaches. In contrast to the model by Heinzel et al. (2019) and the clinical model, the digital risk score assigned similarly high probabilities of having PD to several individuals in the prodromal group as to individuals with diagnosed PD. Six individuals in the prodromal group converted to PD before the digital sensor data was collected with a mean time since diagnosis of 2.14 ± 1.6 years, compared to 7.18 ± 2.02 years for the diagnosed PD group. Across all models these individuals who were initially recruited to the prodromal group and were subsequently diagnosed with PD were assigned high risk scores, except one, demonstrating that all models can also identify those in earlier disease stages.

**Figure 4:**
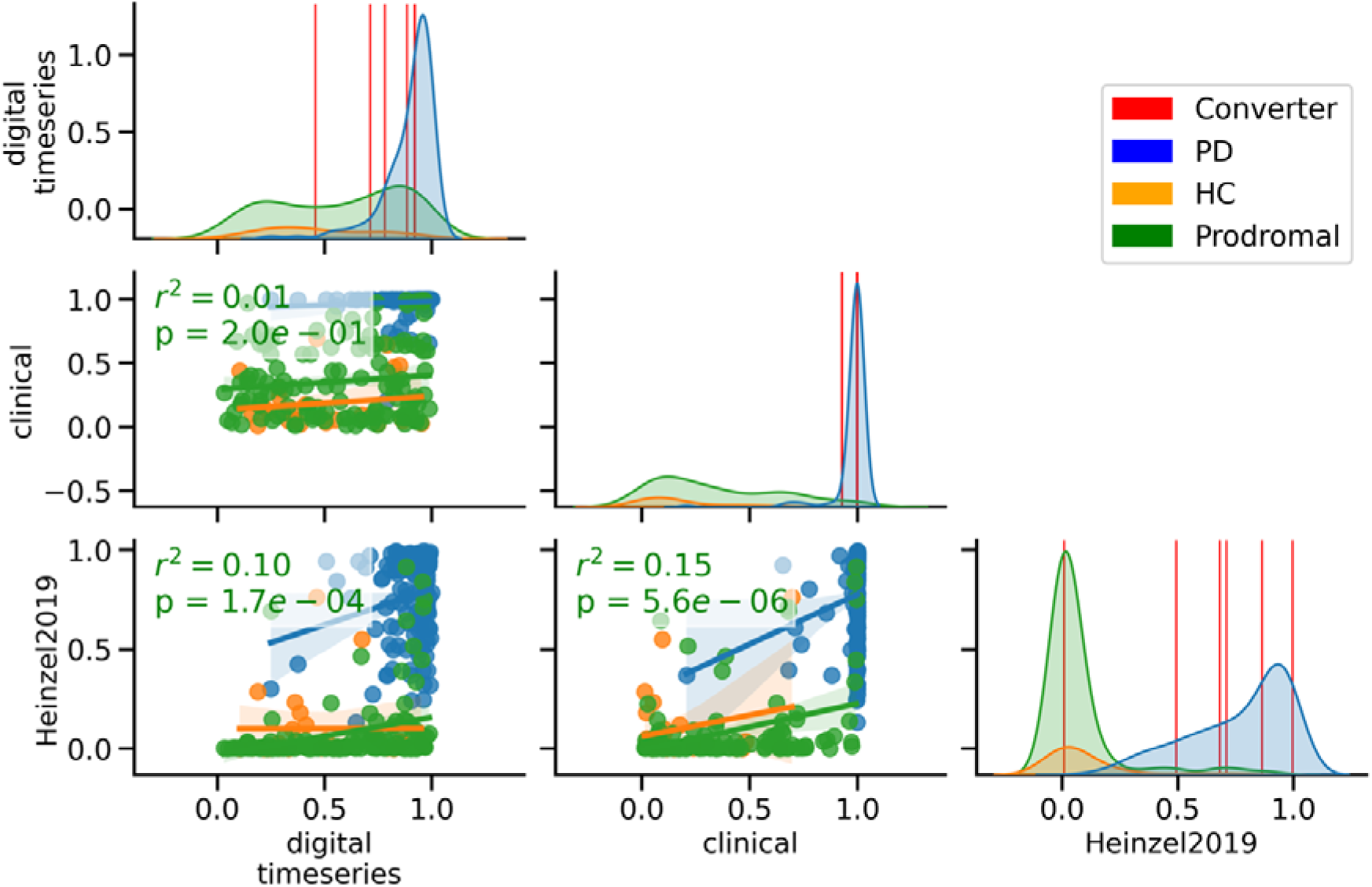
Relation of predicted risk probabilities across models. The plots on the diagonal show the distribution of predicted probabilities of each group for each model. The red lines represent the six participants recruited to the prodromal group who converted before the smartwatch data was collected (overlaps lead to less than six lines appearing on the plots). The scatterplots display for each group the relation between the pairs of models with the coloured lines indicating their linear relationship. The green text notes the r-squared and the p-value of the linear relationship for the prodromal group.

We extracted those individuals who were assigned the highest risks and thus were identified as probable PD according to each model and computed the overlap between methods. All the individuals assigned as probable prodromal PD by the published calculators were in the highest risk group in the digital models (see Methods, Supplemental Table 8). Overall, the digital risk score identified more people as high risk than the established model (76 compared to 2), with only a small overlap between people identified as high risk by the digital model and the clinical model (62.5% of clinical in digital, 34.7% of digital in clinical), thus showing higher sensitivity.

We investigated which known risk factors and prodromal symptoms implied significant differences in risk scores. We found that individuals who had subthreshold parkinsonism (UPDRS III > 6), hyposmia, cognitive impairment, erectile dysfunction, or depression were assigned higher risk scores (Figure 5, Supplemental Table 9). The prodromal risk score was derived from these risk factors and prodromal symptoms, hence serving as a validation. The clinical risk score was built using information on cognitive tests and UPDRS scores which were used to inform the prodromal symptoms of cognitive impairment and subthreshold parkinsonism, thus creating circularity. The digital risk scores, despite not being built with this data, captured significant differences for subthreshold parkinsonism (FDR corrected p-value = 1.33×10^-5^) and depression (FDR corrected p-value = 3.95×10^-2^). Suggestive differences for hyposmia were found as well (uncorrected p-value = 3.63×10^-2^) Of note, individuals with a family history of PD were not assigned significantly higher scores but lower ones, even in the established models, where this was an included risk factor.

**Figure 5:**
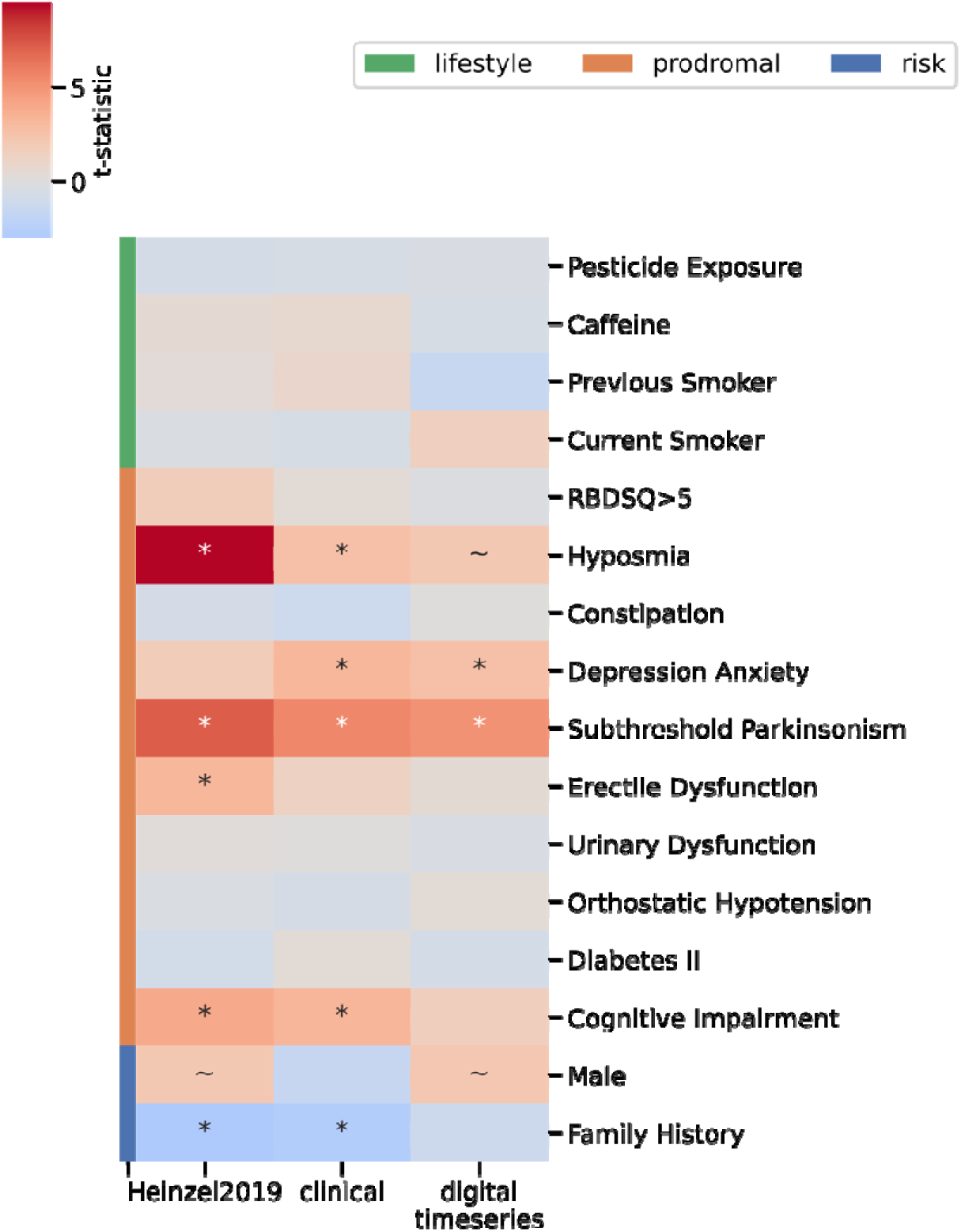
Digital risk score captures known prodromal risk factors. The heatmap shows the difference in predicted probability between at risk and not at-risk individuals (y-axis) for each model (x-axis). The t-statistic coefficient is displayed on a divergent colormap with red indicating at-risk individuals having higher risk scores and blue at-risk individuals having lower risk scores. An asterisk indicates if the 0.05 FDR corrected significance was reached, and a tilde indicates suggestive importance at 0.05.

## Discussion

Here, we leveraged the long-term continuously collected smartwatch data from the PPMI cohort to derive digital timeseries features capable of informing clinical and biological progression, disease subtypes, and risk of being diagnosed with PD.

These digital timeseries components represented the rate of progression in several clinical domains, including motor symptoms and signs, independence in daily living, medication requirements and complications, and autonomic features. One digital timeseries component also related to the rate of ptau and ttau level progression since diagnosis. In addition, PD clusters derived from the digital timeseries data revealed distinct subtypes from those derived with clinical data and were most similar to biological clusters. Finally, we showed that individuals at high risk according to risk factors and prodromal markers were also assigned high risk scores by a model based on digital timeseries features.

Previous work relating digital features to PD progression is limited. Our finding that digital timeseries components representing pulse rate and RMSSD correlate with the rate of progression of motor signs and symptoms, and independence in daily living could represent the known association between self-reported higher physical activity and slower decline in motor and cognitive function as well as slower decline in the ability to perform activities of daily living (ADL) (Amara et al., 2019). In general, higher physical activity has been linked to better cognitive outcomes in PD (Ahlskog, 2011). To our knowledge no previous studies have reported a link between digital markers and tau progression although sleep disturbances and tau pathology have been linked before (Cantero et al., 2010; Holth et al., 2019). The digital timeseries component representing tau progression had WASO and NREM sleep time as representative features, confirming the relation of tau and sleep.

Previous work on sub-typing has been limited to clinical and biological data gathered at clinical visits and has not previously considered digital data continuously collected at-home. We showed that though digital data did not lead to the same subtypes as those that have been previously published (Dadu et al., 2022; Fereshtehnejad et al., 2017; Zhang et al., 2019) they did more closely resemble a clustering derived from biological data than any previously published clustering. Clinical clusters not reflecting biological subtypes potentially indicate a lack of concordance with the underlying pathophysiology, and thus may not be well suited to informing treatment response.

We have previously demonstrated the ability to identify those who will go on to receive a future a diagnosis of PD using a single week of accelerometer sensor data (Schalkamp et. al, 2023). Here, we evaluated long-term digital markers derived from a multi-sensor device worn by individuals harbouring risk genetic variants or prodromal markers for PD. By highlighting the sensitivity of the digital risk score to identify all individuals with probable prodromal PD as calculated with established risk scores (Heinzel et al., 2019), we further highlight the potential of digital devices for early screening and risk stratification.

The primary limitations of this study relate to data availability and choice of methodology. Due to the Verily Study Watch only being introduced 10 years after the start of the PPMI study, the temporal overlap between the clinical assessments and digital data was limited. PPMI recruited *denovo* PD cases in 2008 which, by the time of Verily Study Watch data collection, had already been diagnosed for several years with the majority prescribed symptomatic medication. Conclusions about early disease monitoring with digital devices can therefore not be determined from our current study. For the comparison of sub-typings, we limited our study by only including three published clusterings while several other options exist (Mestre et al., 2021). Further, the considered clusterings were each trained on different subsets of subjects and data collected at different times, for example our clinical clustering considered all clinic visits between 2010 and 2021 whereas the digital clustering only considered data collected between 2018 and 2022. The included previously published sub-typings further applied their own restrictions on subject and data inclusion, thus again leading to a different dataset considered. The resulting overlap between all clusterings was thus small. Regarding the risk calculations, the prodromal group in PPMI only included 28 known converters of which only seven had digital data available with six converting before data collection and one without a known conversion date, limiting the assessment of the true risk of developing PD. Our study was further limited to the derived features provided by Verily, the code for which is proprietary, limiting reproducibility in other cohorts.

In conclusion, long-term digital monitoring can provide valuable insights into the disease course and the heterogeneity of PD via monitoring during the diseased stage but can also inform disease risk via monitoring of people at-risk.

## Supporting information

All Supplemental Tables

## Data Availability

All data used in the present study are available online under https://www.ppmi-info.org/access-data-specimens/download-data.

## Data Availability

All data used in this study was accessed from https://www.ppmi-info.org/access-data-specimens/download-data. The Institutional Review Board approved the PPMI program and all participants gave written informed consent. For up-to-date information on the study, visit www.ppmi-info.org.

## Code Availability

All associated code to reproduce the analyses performed here will be made publicly available upon publication.

## Acknowledgements

We thank all participants of the PPMI study, all investigators, and the Michael J. Fox Foundation.

## Author Contribution

A-K.S. and C.S. participated in designing the study, topic definition, and review of relevant studies. Machine learning models and statistical analyses were designed and implemented by A-K.S.. Figures and tables were done by A-K.S. with the support of C.S.. A-K.S. wrote the first draft. A-K.S., C.S., N.A.H., K.J.P., and V.E.-P. contributed to subsequent versions of the manuscript. All authors critically reviewed the paper, all authors have a clear understanding of the content, results, and conclusions of the study and agree to submit this manuscript for publication. The corresponding author (C.S.) declares that all authors listed meet the authorship criteria and that no other authors involved in this study are omitted. C.S. is ultimately responsible for this article.

## Methods

All analyses were performed in python v3.9 using sklearn 1.2.1 (Pedregosa et al., 2011) for model training and evaluation, tsfresh 0.20.0 (Christ, 2018) for timeseries feature extraction, scipy 1.10.0 and pingouin 0.5.3 for statistical testing (Vallat, 2018), and matplotlib 3.6.3 and seaborn 0.12.2 for creating figures. Data loading and manipulation has been facilitated through an adapted version of pypmi (https://github.com/rmarkello/pypmi). All associated code will be made available at (https://github.com/aschalkamp/DigitalPPMI).

### Study Cohort

The Parkinson’s disease progression marker initiative (PPMI) has collected data from those with recently diagnosed (*de novo*) PD, people at risk, and unaffected controls since 2010. They put a focus on longitudinal data collection of brain imaging, blood, urine, CSF, and behavioural data. Since 2018 a subset of participants has been supplied with a Verily Study Watch, which is equipped with a multitude of sensors including accelerometer, gyroscope, electroencephalography (ECG), and photoplethysmography (PPG). We used the analytic dataset cohort assignment, which provides the most up-to-date assignment of subjects into PD, healthy control, Scans Without evidence of dopaminergic deficit (SWEDD), and prodromal class. The derived digital marker data was available for 149 PD cases, 158 prodromal cases, and 35 healthy controls. The prodromal group in this study was formed of people at high risk of PD, namely the recruiting criteria were rapid eye movement behavioural sleep disorder (RBD), olfactory loss, or mutations in Mendelian inherited genes considered causative or contribute to increased risk in PD (e.g. *LRRK2, GBA, SNCA, Parkin, Pink1*) with or without dopamine transporter (DAT) deficiency. Seven individuals received subsequently a diagnosis of PD, six before smartwatch data collection, for one the date of diagnosis was unknown. PD cases were no longer than two years post diagnosis, and unaffected controls were shown to demonstrate no visual deficit on DaTScan.

#### Digital Data

From the raw Verily Study Watch data several derived measures were provided by Verily (Supplemental Table 1) and were accessed by us through the LONI website of PPMI in November 2022. These included 1-hour interval timeseries data on sleep, physical activity, and vital signs. Each derived digital marker had a different availability. The hourly step count data covered an average of 1.25 years (std = 0.54) with a mean recorded time of 0.91 years (std = 0.52). The time between the measurements ranged from one hour to 6.76 days with a mean of 137.56 minutes (std = 530.76). The sleep data covered an average of 1.19 years (std = 0.57) with a mean covered time of 8.4 days (std = 6.68). Here, the time between measurements was larger on average with 6.35 days (std = 19.83).

### Clinical and biological data

Data was downloaded from LONI PPMI in 2021. The following clinical assessments were retrieved: motor assessments including UPDRS scores (part II, III) (Goetz et al., 2008), cognitive assessments like Montreal Cognitive Assessment (MoCA) (Nasreddine et al., 2005), semantic fluency, Benton judgement of line orientation (Benton et al., 1978), and WMS-III Letter-Number Sequencing Test (LNS), Hopkins Verbal Learning Test (HVLT) (Shapiro et al., 1999), and symbol digit, psychiatric questionnaires including State-trait-anxiety inventory (STAI) (Spielberger, 1983), geriatric depression score (GDS) (Sheikh, 1986), and Impulsive-Compulsive Disorders in PD (QUIP) (Weintraub et al., 2009), as well as autonomic assessments like Scale for Outcomes in Parkinson’s disease for Autonomic Symptoms (SCOPA) (Visser et al., 2004), Epworth Sleepiness Score (ESS) (Johns, 1991), REM sleep behavior disorder screening questionnaire (RBDSQ) (Stiasny-Kolster et al., 2007), and systolic blood pressure drop, assessments of daily functioning with the modified Schwab England Activities of Daily Living (ADL) and the UPDRS I, and medication information (UPDRS III ON, UPDRS IV, Levodopa Equivalent Daily Dosage (LEDD)). The six prodromal cases who converted to PD before digital data collection also converted before their respective last available visit to the clinic.

The imaging biomarker data was calculated from DaTscan imaging with striatal binding ratio (SBR) for caudate and putamen for each hemisphere being available via LONI. From these four measures we computed the overall mean SBR, the mean SBR for each hemisphere, the mean SBR for putamen and caudate, the SBR asymmetry overall and for caudate and putamen, and the minimum putamen SBR. We considered the following biospecimen data: total tau (ttau), phosphorelated tau (ptau), total alpha-synuclein, Amyloidbeta 42-1, Glial Fibrillary Acidic Protein (GFAP), and Neurofilament Light chain (NFL).

These data were collected, cleaned, and merged based on the subject identifier and visit date. For UPDRS III we distinguished between ON and OFF assessments with OFF being those where the subject was not on medication either due to not taking medication at all or because the medication was deliberately not taken for this assessment, and ON being all assessments conducted when the subject took their medication.

### Prodromal markers and risk factors

We further retrieved all data necessary to calculate the prodromal risk score as outlined in Heinzel et al. (2019) (Supplemental Table 7). This included the risk factors: age, sex, exposure to pesticides, caffeine usage, smoking history, family history or genetic markers, diagnosis of diabetes mellitus type II. And the prodromal markers: RBD, positive DaTscan, subthreshold parkinsonism, hyposmia, constipation, daytime somnolence, orthostatic hypotension, urinary dysfunction, depression, and global cognitive deficit. We were not able to retrieve information from PPMI corresponding to self-reported physical activity, substantia nigra hyperechogeneicity, and urate levels as detailed in the criteria. We decided to include family history information over genetic data as described in the model.

### Covariate Adjustment

The overall mean over time was computed for each subject for each digital marker. The PD group was significantly younger than the prodromal group (t = −7.89, p-value = 6.06×10^-14^) thus linear models were fit for each digital marker on the healthy controls (N = 35) to identify the effect of age and sex on each marker. The residuals were then compared between groups with two-sided T-tests.

### Progression estimates

The longitudinal data of each data modality was analysed separately to retrieve estimates of progression.

### Clinical and Biological

The progression of the clinical and biological data was modelled for the PD group with linear mixed models using the time of diagnosis as the origin. The clinical measures were transformed, if needed, such that higher scores represented more impairment. Random intercepts and slopes were fitted to allow for variation of the individuals around the population mean. The intercept was interpreted as the severity at diagnosis and the slope as the speed of progression. We restricted the data for each modality individually to subjects with at least two visits to retain the most data possible. Overall, this data was available for 665 PD cases with diagnosis age of 60.24±10.10 years. They were followed for an average of 5.91±2.77 years with 11.33±4.66 visits which were on average 208±130 days apart. We computed principal components via Principal Component Analysis (PCA) for the clinical intercepts and slopes separately for defined modalities: motor (UPDRS III OFF, UPDRS II), daily (Schwab England ADL, UPDRS I), cognitive (MoCA, Benton, LNS, HVTL recall, retention, and recognition, symbol digit, semantic fluency), psychological (STAI trait, STAI state, GDS, QUIP), autonomic (ESS, RBDSQ, SCOPA autonome, systolic blood pressure drop), DaTscan (mean SBR, putamen SBR, caudate SBR, asymmetry, caudate asymmetry, putamen asymmetry), Amyloidbeta, ttau, ptau, alpha-synuclein, GFAP, NFL. The same method was used for the medication domain (UPDRS III ON, UPDRS III OFF, UPDRS IV, LEDD) but the time since first medication was modelled instead of time since diagnosis. The principal components were the identified based on UPDRS III ON-OFF, UPDRS IV, and LEDD. On average the first principal component explained 0.76±0.25 of the variance. The minimum explained variance ratio of the first principal component was 0.41 for medication severity.

### Digital

Tsfresh was applied for each individual for each digital feature to extract their timeseries features. This included 783 derived features like the maximum, minimum, skewness, kurtosis, and trend for each timeseries. The method does not consider the time intervals between data, leading to potential biases where data was only sporadically worn. We ran PCA on all digital timeseries features from tsfresh. The first 10 principal components explained 27% of the total variance.

### Correlation of progression estimates

We aimed to study the correlation of the progression estimates of the different data modalities. The principal components of the clinical features were correlated with the principal components of the digital features via Pearson correlation. For those principal components that were significantly correlated with clinical features, we extracted the PC loadings, representing the correlation of each timeseries feature with that principal component (Supplemental Figure 3).

### Sub-typing

#### Replicating clinical subtypes from literature

We replicated one published sub-typing method and retrieved the public sub-typing data of two others. Dadu et al. (2022) and Zhang et al. (2019) published the clusters for the PPMI subjects, which we retrieved. We repeated the analysis steps outlined in Fereshtehnejad et al. (2017) to retrieve similar clusters as they suggested for the last clinical visit available.

### Own clusters

We retrieved subtypes based on digital and clinical data respectively using our own method. We made use of the retrieved progression estimates and applied six different clustering techniques: kmeans, gaussian mixture, hierarchical Ward linkage, spectral clustering, agglomerative clustering, and Birch. We decided to retrieve three subtypes to allow for comparability to the existing methods as they all report three subtypes. The best clustering method was evaluated based on silhouette score. Ward clustering (silhouette = 0.25) was identified as the best (Supplemental Table 10). We identified the clusters for a total of four datasets: all timeseries features, the clinical severity and progression estimates, and the CSF severity and progression estimates, and the DaTscan severity and progression estimates.

### Comparison of clusters

We evaluated the similarity between the clusters using a composite score of three metrics that are invariant to label permutations: Fowlkes-mallow (Fowlkes & Mallows, 1983), normalised mutual information (Vinh et al., 2009), and adjusted rand score (Hubert & Arabie, 1985). The adjusted rand score ranging from −1 to 1 was rescaled to 0-1 to have the same range as the other metrics. These metrics were computed for each pair of clustering methods and the mean over these metrics was reported (Supplemental Table 5).

### Significant differences between clusters

We evaluated for all principal components of the progression estimates from the digital, clinical, and biological data, how the identified clusters differed. We did so by computing two-sided T-tests between each pair of clusters for each clustering method.

### Parkinson’s disease risk

The reporting on the comparison between risks was performed on the intersection between the datasets. 25 subjects recruited to the prodromal group converted with seven of those participating in the smartwatch study. We reported on these converters separately.

### Published risk scores

The model described in Heinzel et al. (2019) was implemented. First, the prior was retrieved for each subject based on the age. Then the likelihood ratio was computed by multiplying all increasing and decreasing factors related to the measures listed in Supplemental Table 7. This likelihood was then converted to a probability: odds = ((prior/100)/(1-(prior/100))) * ‘LR, probability = odds/(1+odds).

### Own predicted risk

We retrieved the predicted risk by training lasso logistic regression models identifying PD from healthy controls. The classifiers were trained by first selecting relevant features for the binary distinction with tsfresh “select_features”. A nested cross-validation was used with an inner and outer five-fold stratified split. Based on the training set extracted with the outer fold split, the data was standardised. The inner split was used to run a grid search to identify the best lasso penalty parameter between 10^-2^ and 10^2^. The area under the precision recall curve (AUPRC) was used as the evaluation score and the final model comparison. We trained models on two different sets of predictors: last available visit to the clinic (MoCA, Benton, LNS, HVLT recall, HVLT recognition, HVLT retention, semantic fluency, symbol digit, QUIP, GDS, STAI trait, STAI state, SCOPA autonome, ESS, RBDSQ, systolic blood pressure drop, Schwab England ADL, UPDRS I, UPDRS II, UPDRS III OFF) and tsfresh extracted timeseries features. The predictors were assessed for stability and significance across folds.

The predicted probabilities were retrieved for all subjects, including the external test set of prodromal, as the average over the outer folds.

Digital timeseries data achieved high, but significantly lower, accuracy in identifying PD from healthy controls than models trained on the most recent clinical assessments (Supplemental Figure 4). The most stable feature in the clinical model was UPDRS II (p-value = 4.36×10^-4^). Contributing factors in the digital model were the skewness of total REM sleep time (p-value = 4×10^-6^) and hourly step count sum linear trend (p-value = 4.7×10^-5^).

### Comparison of predicted risks

We compared these predicted probabilities with the prodromal risk score calculated using Heinzel et al. (2019). We did so by correlating each pair of predicted probabilities. We further assessed which known prodromal markers and risk factors were associated with higher predicted probabilities for each classifier using two-sided T-tests. Finally, we compared the identified high-risk individuals of each modality by calculating the percentage of overlap and cohen’s kappa.

