## Supplemental Figures, Tables, Legends for "Leveraging long-term smartwatch data to inform Parkinson’s disease progression, subtypes, and risk"

#### Supplements

Supplemental Table 1: Derived digital markers as provided by Verily

The different hourly statistics as derived from the smartwatch data are described.

This information is taken from the accompanying documents on PPMI LONI.

| modality | category | sensors | #features | features | model |
|---|---|---|---|---|---|
| physical activity | ambulatory | 3-axis accelerometer | 1 | hourly ambulatory minutes | 2-class classifier (walk/run vs other) trained on 215000 hours of self-report labelled free-living data from 1800 adult subjects with out-of sample performance of 87% |
| | step | 3-axis accelerometer | 1 | hourly step count | frequency-based model validated against ankle-worn gait monitor on 329 days of free-living data of 75 adult subjects with 18% mean absolute error |
| sleep | sleep onset/offset | accelerometer, PSG | 4 | sleep efficiency, number of awakenings, total sleep time, wake after sleep onset | algorithm trained on PSG and ECG validated against majority vote of three wearables on 176 nights in home setting of 50 adult subjects with median absolute error of sleep onset of 6 minutes and 9 minutes for sleep offset |
| | sleep stages | accelerometer, PSG | 4 | REM, NREM, light NREM, deep NREM | algorithm trained on PSG and ECG validated against majority vote of three wearables on 176 nights in home setting of 50 adult subjects with an overall accuracy of 70% (Sridhar et al., 2020) |
| vital signs | pulse rate | PSG | 1 | total mean pulse rate per hour | algorithm from ADI validated against heart rate of ECG on one to two hours of in-clinic data of 50 adult subjects with a mean |

| | | | | |
|---|---|---|---|---|
| | | | | absolute error of 10.7 beats per minute ADI2023 |
| pulse rate variabilit y | PSG | 3 | mean, median and variance of hourly RMSSD | algorithm measuring RMSSD of interbeat intervals validated against 510000 wearables for ECG on 200 days of free-living data of 50 adult subjects with mean absolute error of 9.9ms (Billman, 2011) |

Supplemental Table 2: Significant differences in digital markers between groups

The mean value per residual mean digital marker corrected for age and sex is shown for the healthy controls, prodromal cases, and the diagnosed Parkinson’s disease cases together with the sample size per group. We show the statistics of the two sided T-test as the t-statistic and p-value. The digital markers are here the mean over the whole observation time for each individual.

Supplemental Table 3: Significant differences in digital markers between groups

The mean value per residual mean digital marker corrected for age and sex is shown for the healthy controls, the diagnosed PD, and the prodromal GBA, LRRK2, SNCA, hyposmia, RBD, and DaTscan positive cases together with the sample size per group. We show the statistics of the two sided T-test as the t-statistic and p-value. The digital markers are here the mean over the whole observation time for each individual.

Supplemental Table 4: Correlation between clinical and biological severity and progression with digital timeseries features

For the diagnosed PD cases we show the statistics of the pearson r correlation as the r statistic, p-value, and number of samples for each pair of progression estimate and severity value with the first 10 principal components of the digital timeseries features.

Supplemental Table 5: Similarity between clusters

For each pair of clusters we display the four similarity metrics and the composite overall score.

Supplemental Table 6: Cluster differences between progression estimates

For each cluster method we show the differences between the clusters in terms of the progression estimates for clinical, biological, and digital features. The statistics of the two sided T-test are shown as the t-statistic, p-value, number of samples, FDR corrected p-value, and significance.

Supplemental Table 7: Risk factors and Prodromal Markers

We describe the process of obtaining risk and prodromal markers from PPMI data. The selection of markers was taken from Heinzel et al. (2019).

| | | |
|---|---|---|
| risk factors | | |
| | age | age at data retrieval date: 01.10.2021 |
| | sex | male |
| | pesticide exposure | FOUND questionnaire whether occupational exposure |
| | non-use of caffeine | FOUND questionnaire less than 6 cups of tea or 3 cups of coffee weekly |
| | never smoke | FOUND questionnaire not ever smoked regularly |

| | | |
|---|---|---|
| | previous smoke | FOUND questionnaire ever smoked regularly and not smoke currently |
| | current smoke | FOUND current regular smoker |
| | 1st degree relative with PD | mother, father, or sibling with PD diagnosis |
| | diabetes mellitus type II | medical condition log searched for '(?!.*pre)(?!.*borderline)((.*(II|2|two).*Diabet.*)|.*Diabet.*type.*(II|2|two).*)' |
| prodromal markers | | |
| | proven RBD | medical condition log searched for '.*(REM behavi|RBD|Rapid Eye).*' or listed under confirmed RBD in analytic dataset |
| | RBD test | ever scored higher than 5 on RBDSQ |
| | positive DaTscan | visual inspection of DaTscan abnormal or minimum putamen SBR below 60th precentile |
| | subthreshold parkinsonism | ever UPDRS III score above 6 |
| | olfactory loss | medical condition log searched for '.*(hyposmia|anosmia).*' or listed under confirmed hyposmia in analytic dataset or ever scored below 1.5 std from age and sex matched mean (Brumm et al., 2023) |
| | constipation | medical condition log searched for '.*constipation.*' |
| | excessive daytime sleepiness | medical condition log searched for '.*sleepiness.*' |
| | urinary dysfunction | medical condition log searched for '(?!fecal).*incontinence.*' |
| | orthostatic hypotension | medical condition log searched for '.*hypotension.*' |
| | erectile dysfunction | medical condition log searched for '.*erectile.*' |
| | depression | medical condition log searched for '.*(anxiety|depression).*' |
| | cognitive deficit | ever cognitive categorisation listed as mild impairment or dementia |

Supplemental Table 8: Overlap between identified high-risk individuals

For each pair of risk models, we report the cohen's kappa and the percentage of overlap defined as intersection/N_highrisk_modality1 and intersection/N_highrisk_modality2.

Supplemental Table 9: Differences in risk scores between risk factor and prodromal symptom carriers

The results of the two sided T-tests are shown as the t-statistic, p-value, and number of individuals for each pair of risk factor/prodromal symptom and risk score.

Supplemental Table 10: Silhouette Scores for different clustering methods

For each clustering method the silhouette score is shown for the derived digital clusters.

Supplemental Figure 1: Mean digital markers are affected in people diagnosed with PD

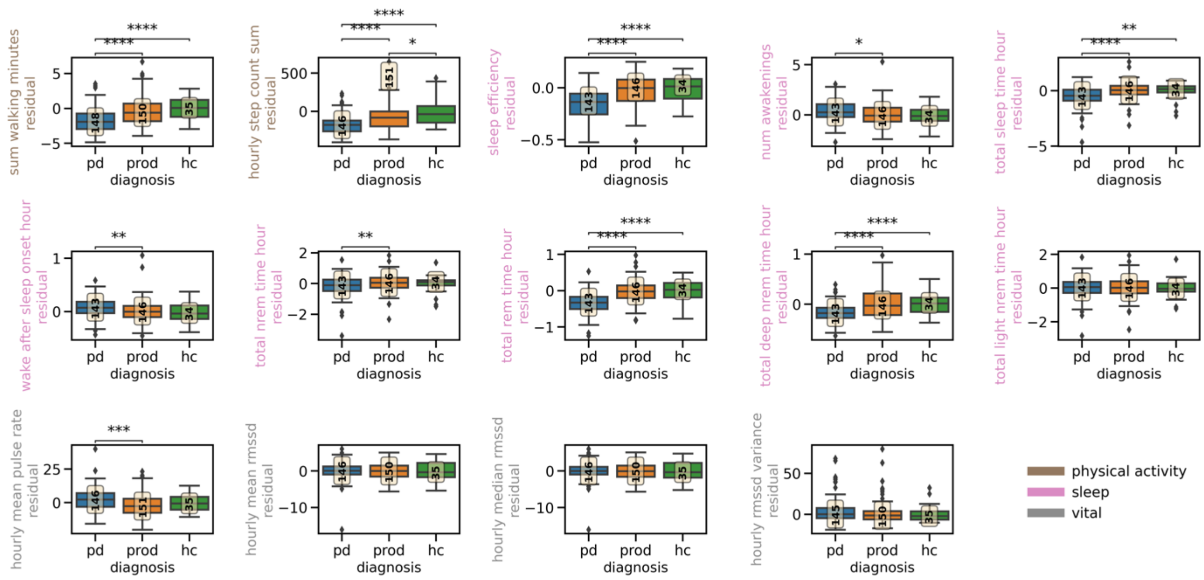


Boxplots show the residual overall mean over the whole observation time per subject for each group for each digital marker. The boxplot depicts the group mean and quartiles per diagnosis group (Parkinson’s disease cases:pd; prodromal cases (higher risk group, converters removed): prod; and healthy controls:hc). The number in the yellow box indicates the number of individuals per group. Group differences calculated with two-sided T-test after 0.05 Bonferroni-correction are shown with asterisk. No indication/not significant: 1.67e-02 < p <= 1.00e+00, *: 2.00e-03 < p <= 1.67e-02, **: 2.00e-04 < p <= 2.00e-03, ***: 2.00e-06 < p <= 2.00e-04, ****: p <= 2.00e-06.

### Supplemental Figure 2: Mean digital markers across prodromal risk groups

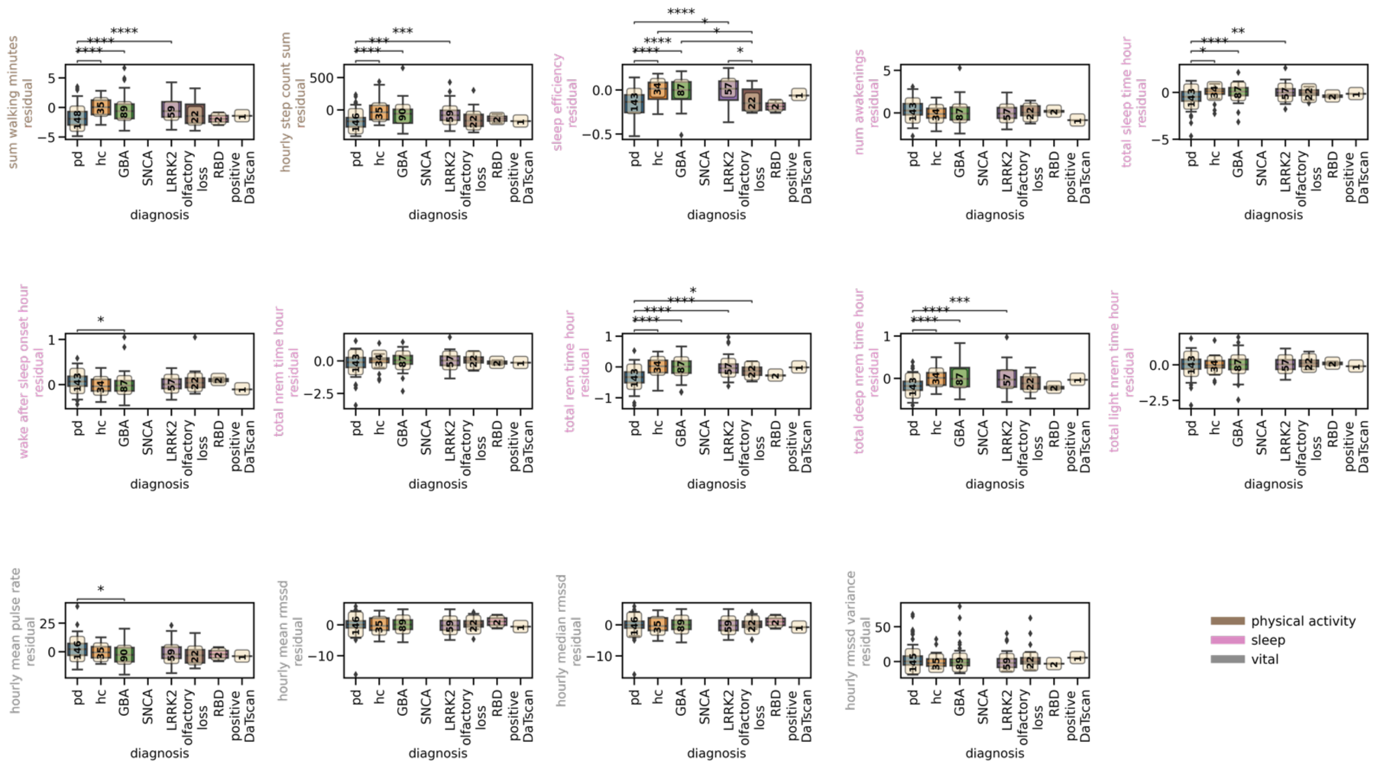


Boxplots show the residual overall mean over the whole observation time per subject for each group for each digital marker. The boxplot depicts the group mean and quartiles per group (Parkinson's disease cases: pd; healthy controls: hc; carriers of genetic risk alleles or prodromal symptoms without a diagnosis of PD: *GBA, SNCA, LRRK2*, olfactory loss, RBD, positive DaTscan). The number in the yellow box indicates the number of individuals per group. Group differences calculated with two-sided T-test after 0.05 Bonferroni-correction are shown with asterisk.

Supplemental Figure 3: PCA loadings for the significant digital timeseries components

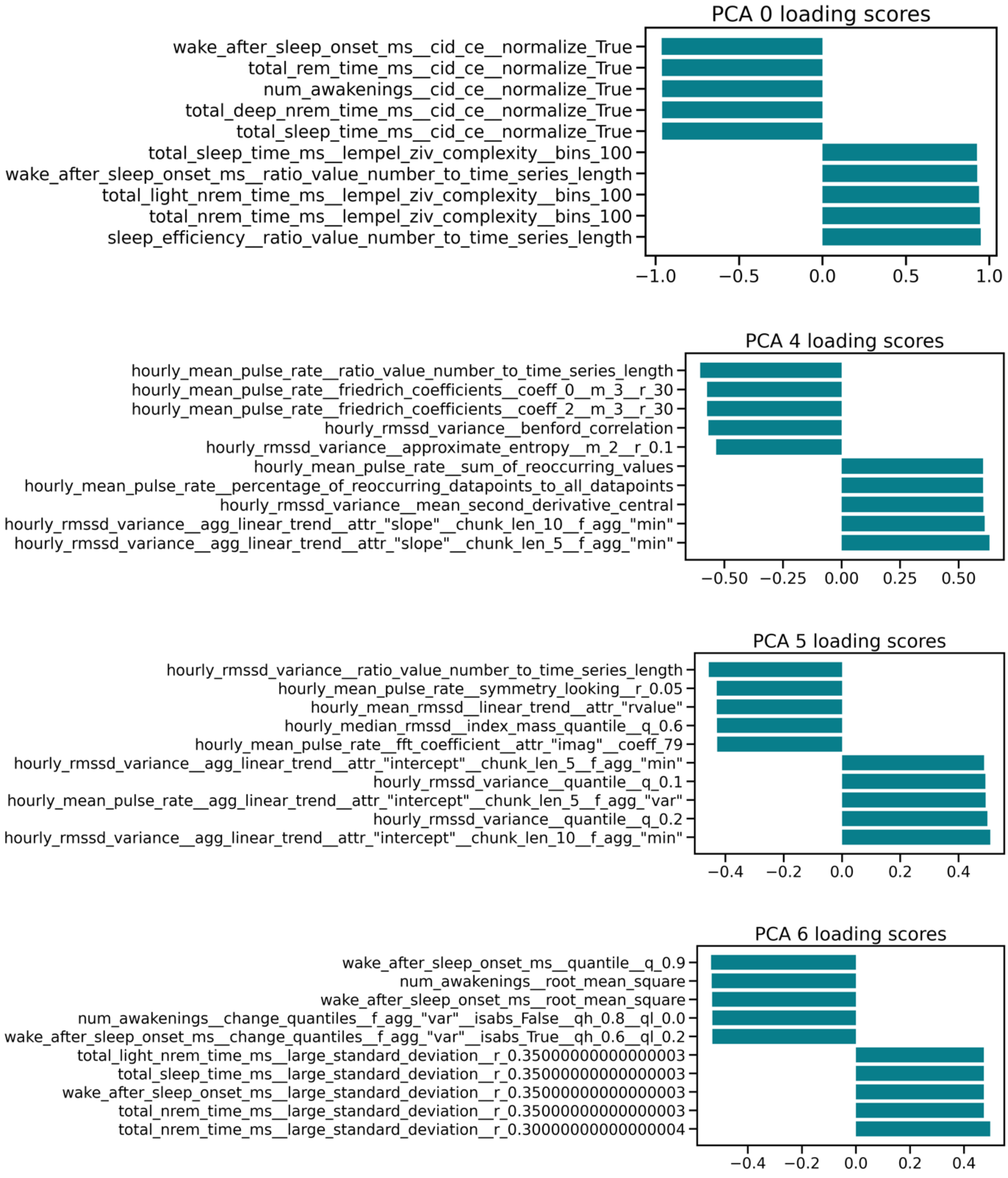

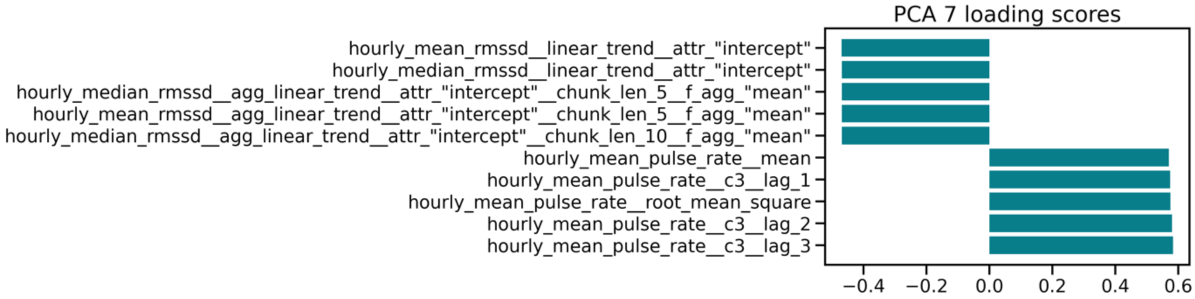


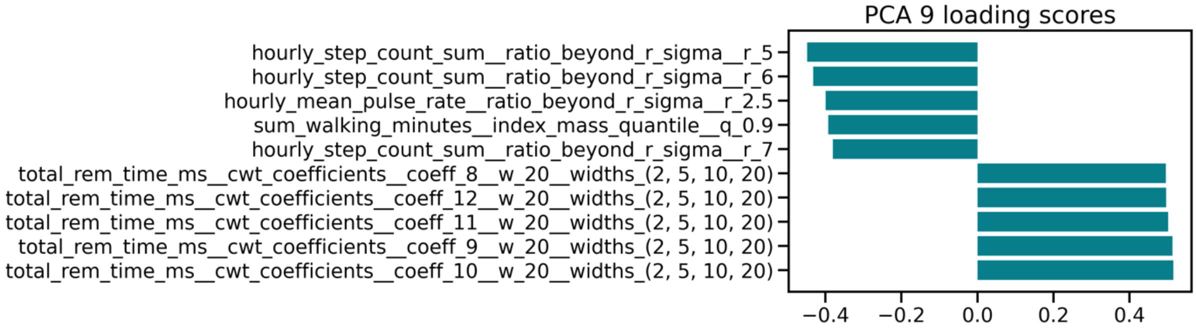


For the principal components 0, 4, 5, 6, 7, and 9 the top and bottom five features with the largest effect sizes are displayed with the bar plot indicating the direction of effect and effect size.

#### Supplemental Figure 4: Model performance identifying Parkinson's cases from healthy controls

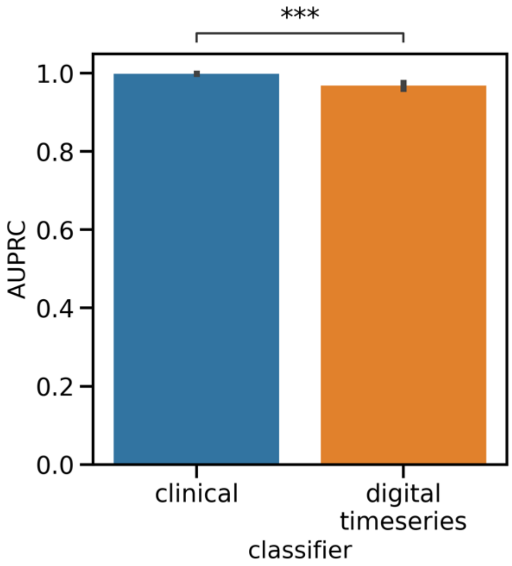


The mean AUPRC is shown across the outer five folds of the cross-validation with the errorbars indicating the 95% CI. The asterisks indicate significant differences in performance assessed with two-sided T-tests.
